# Community Knowledge, Perceptions and Attitudes Regarding Buruli Ulcer in Bafia Health District, Centre Region-Cameroon

**DOI:** 10.1101/2025.08.14.25333658

**Authors:** Earnest Njih Tabah, Chefor Alain Djam, Irine Ngani Njih, Loic Douanla Pagning, Colin Tsago Nzoyem, Elisabeth Baran-A-Bidias, Christian Kouayep-Watat, Franck Eric Wanda, Larraitz Ventoso

**Affiliations:** National Yaws, Leishmaniasis, Leprosy and Buruli ulcer Control Programme, Ministry of Public Health, Yaounde, Cameroon; Department of Public Health, Faculty of Medicine and Pharmaceutical Sciences, University of Dschang, West Region, Cameroon; Research Initiative in Tropical and Community Health (RITCH), Yaounde, Cameroon; FAIRMED – Cameroon. P.O. BOX 5807 Yaounde, Cameroon; Department of International Cooperation, ANESVAD Foundation, Henao, 29-31 (oficinas) 1° planta 48009 BILBAO, Spain

**Keywords:** Community, Knowledge, perceptions, attitude, and Buruli Ulcer disease

## Abstract

**Background:** Buruli ulcer (BU), a neglected tropical disease, occurs in about thirty-three countries tropical countries world-wide. Misconceptions about BU leads to poor health-seeking behaviors. This study explored community knowledge, perceptions and attitudes regarding BU.

**Methods:** We conducted a cross-sectional household survey in selected communities of Bafia Health District, Centre region of Cameroon.

**Results:** We had 1,341 respondents aged 10–87 years and a mean age of 34.8±18.4yrs. About 53% were males, 50.1% were married, and majority were Christians. Some 30.1% had heard about and 21.1% knew someone with BU. Major sources of information on BU were: family members(29%), friends(21%) and school(14%). Only 17.2% of respondents correctly identified a manifestation of BU, and 2.3% knew BU its cause. A considerable proportion attributed BU to witchcraft(8%), poor hygiene(9.8%) and spontaneous occurrence(5.3%). Only 19.2% believed BU was curable, of whom 78.6% advised on health facilities, meanwhile 8.2% preferred traditional healers for treatment. Attitudes towards persons with BU (PWBU) were mostly negative as only 27.4% would show them respect, 19,5% would shake hands, and 16.4% would share the same plate with them. Additionally, only 17.3% approved their participation, and 14.1% and 12.2% respectively would allow their child to play with or marry a PWBU. Factors associated with positive attitudes included: having head about BU, knowing a PWBU, understanding that BU is curable and treatable in hospital, having attained at least secondary education. Negative attitudes were associated with beliefs that BU is caused by supernatural forces, poor hygiene, or living with a PWBU.

**Conclusion:** There was poor community knowledge and perceptions about BU in the BHD, which negatively influenced community attitudes towards PWBU. A community education intervention focusing on the natural occurrence, biological etiology, non-hereditary nature, the non-human-to-human transmission, and the curable nature of BU could improve upon the situation in BHD.

**Author summary:** Buruli Ulcer (BU), a neglected tropical disease, leads to soft-tissue destruction and physical deformities if not detected early and treated adequately. Poor community understanding and misconceptions leads to poor health-seeking behaviours among victims. We explored community knowledge, perceptions and attitudes among 1341 participants in Bafia health district, centre region, Cameroon. Our respondents were not familiar and had poor knowledge as well as erroneous perceptions regarding BU. as less than 1/3^rd^ had heard, and less than a quarter knew someone with BU. Less than 1/5^th^ knew the manifestation of BU, and as low as 2.3% knew the its cause, with many attributing BU to witchcraft, poor hygiene and spontaneous occurrence. Very few believed that BU is curable and treatable in health facilities. Attitudes towards PWBU were negative, as very few would shake hands, share the same plate, or allow their participation in community life. Drivers of negative attitudes were the beliefs that BU is caused by supernatural forces, poor hygiene, or living with a PWBU. A community education programmes on BU, focusing on its natural occurrence, the non-human-to-human transmission, and its curable nature, could improve upon community knowledge and attitudes regarding BU in Bafia health district.

## Introduction

Buruli ulcer (BU), a neglected tropical diseases (NTD) is caused by *Mycobacterium ulcerans,* a microorganism belonging to the same genus of bacteria as those responsible for tuberculosis and leprosy, ranking it the third most common mycobacterial infection(1–3). BU occurs in remote areas of tropical Africa, Latin American, Southeast Asia, around aquatic environments characterized by stagnant or slow flowing water bodies or marshy areas(4–6). The exact mode of transmission is still unclear(7,8), although some studies have suggested the involvement of an animal reservoir or of insect vectors(1,9–12). No human-to-human transmission has been described(8). BU affects persons of both gender and of all ages, although children below 15 years of age are more vulnerable compared to adults(13,14). BU disease begins as a non-ulcerative and non-analgesic skin lesion (in the form of papule, nodule, plaque or oedema) after an incubation period of 21 to 90 days. Each lesion ulcerates and progresses to an extensive ulcer, if not detected and treated early(13–16). The extensive ulcers leads deformities, limitation of joint movement and eventually to functional disability, loss of economic productivity (17–20). The deformities and disabilities consequent to BU has led to fear of diseases in endemic communities as well as stimulating erroneous perceptions and poor attitudes towards the BU and those affected by it (19,21,22). Although BU is curable through specific anti-biotherapy associated with limited surgery, physiotherapy, nutritional and psychosocial support(8,23–25), many affected persons of endemic communities still resort to non-convention and traditional remedies as first means of treatment(21,22,26–29). As the exact mode of BU transmission remains elusive(7,8), early detection and adequate treatment are the only measures to prevent damaging consequences of BU(8,23,30,31). Effective implementation of these measures relies on proper mobilization and sensitization of endemic communities; to dissipate the erroneous perceptions and poor attitudes towards BU and increase the uptake of BU control services that are provided to them(15).

Cameroon, has been implementing BU control activities for over 25 years today(15,32). Over this period, new BU endemic health district have continued to emerge(15,33). The Bafia health district(BHD) in the Centre region of Cameroon was recently confirmed endemic for BU, within the frameworks of a project for integrated surveillance of skin neglected tropical diseases (Skin-NTDs) implemented there from 2023 to 2024. As a prelude to extending Buruli ulcer control activities to the district, we embarked on conducting a community-based study to assess the knowledge, perceptions and attitudes regarding BU there.

## Methodology

### Study design

We conducted a cross-sectional descriptive and analytical community-based survey in which a structured questionnaire was administered face-to-face to three people in each household visited to assess knowledge, perceptions and attitudes regarding Buruli ulcer. This study conducted in February 2024, was carried out within the framework of an ANESVAD sponsored project for integrated surveillance of skin-NTDs implemented from August 2023 to November 2024 in Bafia health district of the Centre Region of Cameroon.

### Study setting

The Bafia health district (BHD) is situated some 130 km to the northwest of Yaounde, the capital city of Cameroon (Fig 1). It is found in the country’s central plateau, with dense forest vegetation. The climate is equatorial, with two rainy seasons and two dry seasons. It is crossed by the Mbam and the Sanaga rivers and share borders with seven other health districts. The BHD is made up of 179 villages/communities, distributed into 19 health areas, 16 of which are rural; and had an estimated population 180 733 in 2024(34). The population is constituted of many tribal groups, with the major ones being: the Yambassa, Banen, Bafia, Nyokon, and Yambetta tribes(35).

**Fig 1.**
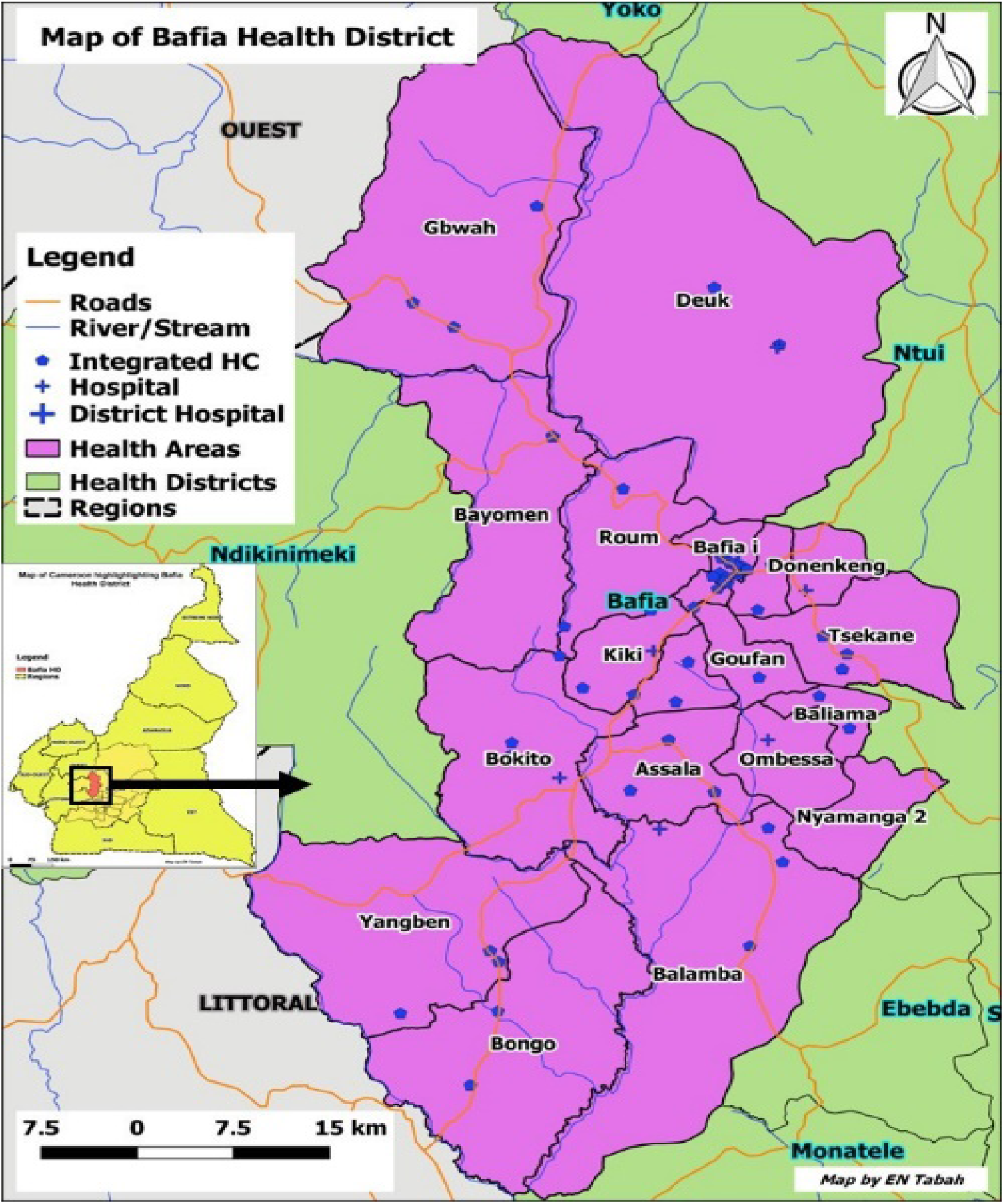
Map of the Bafia Health District (Source: CNLP2LUB-MISANTE)

### Participants, sampling and data collection

#### Participants

We included individuals of both sexes who were 10 years of age and older. People below 10 years of age, community health workers, health personnel and those who did not consent to study were excluded.

#### Sampling

The survey teams visited 3 villages randomly selected from each of the 19 health areas that constitute BHD. In each village, the survey team introduced themselves to the village head, to whom they explained the study objective and procedures, and then sought for his approval to collect data for the study in his village. When approval was granted, the survey teams were then directed to the main square of the village. At this position, they tossed a pencil and then took the street in the direction pointed by the pencil. A systematic random sampling was then used to select households along the street where every fifth household was selected. The immediate next household was visited if a selected household was locked or did not consent to the study. This went on until 10 households were visited in each selected village. In each selected household, an informed consent was sought from the household head or any other adult member of the household present during the visit. The household head and two other people within the household who fulfilled the inclusion criteria and who gave their consent we recruited into the study. The household head gave consent for children under 15 years of age. However, the child-participants assented to participate in the study.

#### Sample size

Based on the assumed proportion of good community knowledge of Buruli ulcer of 84.4% obtained in the southwest of Cameroon(21), and for a 95% confidence interval, an acceptable difference of 0.05 and an estimated non-response rate of 10%, a minimum sample size of 303 was determined for the study.

#### Data collection

Data was collected using a closed ended questionnaire designed to collect demographic data and to evaluate knowledge, perceptions and attitudes regarding Buruli ulcer, adapted from the one we used for KAP-leprosy in Ekondotiti and Mbonge health districts in the Southwest region of Cameroon(36). Excluding demographic data, the questionnaire had 11 questions on knowledge, 6 questions on attitudes and 4 questions on perceptions regarding Buruli ulcer (Table 1).

**Table 1:** List of study variables.

|  | **Variable**<br>**(question)**<br>**Number** | **Variable (Question)** |
| --- | --- | --- |
| **Questions**<br>**on**<br>**Knowledge**<br>**and**<br>**perceptions**<br>**regarding**<br>**Buruli ulcer** | Q1 | Have you heard about Buruli ulcer? |
|  | Q2 | If yes, by which means? |
|  | Q3 | Have you ever seen somebody suffering from Buruli ulcer? |
|  | Q4 | Do you know anybody suffering or who suffered from Buruli ulcer? |
|  | Q5 | Do you have a relative who's suffering or who suffered from Buruli ulcer? |
|  | Q6 | If yes, who is this person to you? |
|  | Q7 | How does Buruli ulcer manifest? |
|  | Q8 | What could be the cause of Buruli ulcer? |
|  | Q9 | If your relative is affected by Buruli ulcer, where would you advise her to go for treatment? |
|  | Q10 | Do you think there is a cure for Buruli ulcer? |
|  | Q11 | What can you do to prevent Buruli ulcer? |
| **Attitudes**<br>**questions** | Q12 | Can you shake hands with somebody suffering from Buruli ulcer? |
|  | Q13 | Can you eat from the same plate with someone who has Buruli ulcer? |
|  | Q14 | Would you be ashamed of a family member suffering from Buruli ulcer? |
|  | Q15 | Would you be willing to let your relative/friend know if you had Buruli ulcer? |
|  | Q16 | Would you let your child play with a person affected by Buruli ulcer? |
|  | Q17 | Would you accept that your child gets married to someone with Buruli ulcer? |
|  | Q18 | Do you think a person with Buruli ulcer should take part in social activities in the community like any other person? |
|  | Q19 | Do you think a person with Buruli ulcer should be employed in the same way as anyone else? |
|  | Q20 | Do you think a person with Buruli ulcer should be respected in the community like any other person? |
|  | Q21 | What kind of difficulties do you think people with Buruli ulcer face in your community? |

Four survey teams of three members each, fluent in the French language were trained on the questionnaire by the principal investigator. The training included full understanding of the questionnaire and techniques of face-to-face interviews. For the practical session of the training, the survey teams were dispatched to four quarters of a semi-urban health area in Yaounde, where they field-tested the questionnaire. Three questions were modified for better understanding after the field-testing exercise. The survey teams were then dispatched to Bafia health district for a period of ten days, where each team covered 4 to 5 health areas and 12 to 15 villages for the data collection phase of the survey.

### Operational definitions

#### High knowledge of Buruli ulcer

A “Yes” answer by 50% and above of the participants to each knowledge question was considered as good community knowledge of regarding Buruli ulcer. Otherwise, it was considered as poor community knowledge

#### Positive attitudes

A “Yes” answer by 50% and above of the participants to each attitude question was considered as positive community attitude regarding Buruli ulcer. Otherwise, it was considered as poor community attitude.

#### Erroneous perceptions

Participants who indicated the cause of Buruli ulcer to be any of the following: curse, divine punishment, witchcraft, heredity, bad food, bad blood; or who believed that BU is not curable; or that victims of BU should not be respected, nor participate in the society nor employed like anyone else, were considered as having erroneous perceptions towards BU

### Ethical considerations

Ethical approval for the study was obtained from the Centre Regional Committee for Research in Human Health, Yaounde, Cameroon (CE N° 0749/ CRERSHC/2023) and administrative authorization from the Regional Delegation of Public Health for the Centre Region. A verbal informed consent was gotten from each participant, after the study objectives and the intended use of the outcome to guide BU control strategies in the Bafia Health District and for publication in scientific journal were explained and they had the chance to ask questions of clarifications. For minors (<18years), their parents consented and then assent sought from them. The verbal consent was preferred when we noted during field testing that participants in remote communities were skeptical about signing or making a thumb print. This procedure was approved by the ethical board if participation was voluntarily. All data were anonymized. There was strict observance of confidentiality in handling and analyzing the data.

### Data management and statistical methods

All filled questionnaires were checked for completeness and correctness and then coded with a unique identifier. The data was then stored in a safe and secure manner until they were analyzed.

Data was entered on Microsoft Excel spreadsheet and cleaned before being exported into IBM SPSS statistics Processor Version 20 for analysis. Proportions were calculated and associations between variables were examined using the Chi-square test, with level of significance set at P-value<0.05. Binary logistic regression using a forward likelihood stepwise entry method was performed by inputting variables that were significant in univariate analyses, to determine predictors of attitudes.

## Results

### Characteristics of respondents

One thousand seven hundred and ten (1710) individuals were approached and 1341 accepted to participate in the study, for a response rate of 78.42%. Their ages ranged from 10 to 87 yrs with a mean age of 34.8±18.4 yrs. There were 708(52.8%) females, and 50.1% were married. Most of the respondents were of the Christian faith, with the most represented being: Catholics (44.7%) and Protestants (29.5%). Muslims constituted 8.4% while animist accounted for 4.2% of the respondents. Majority of the participants were of the Bafia (35.0%) tribe followed by the Yambassa(28.3%), and the smaller tribes regrouped as Mbamois(24.4%) respectively. Participants originating from tribes outside the district accounted for 12.4%. The majority had secondary (54.8%) and primary (35.0%) education levels respectively. The main occupation of the respondents was farming(45.4%) followed by schooling(27.1%).

### Familiarity, knowledge, perceptions regarding BU

The knowledge and familiarity of our respondents regarding BU were generally low as only 30.1% had heard about the disease, 21.1% knew and 16.8% had seen someone with the condition, and about 11% of respondents had relatives with BU(Table 2). For the 404 respondents whom had heard about BU, their sources of information were from family members(29%) friends(21%) school(14%), churches or mosques (5%), Community health workers (5%) and radio or television (5%) (Fig 2). Familiarity with BU was influenced by age, religion occupation and marital status. Older people were most likely to have heard about, knew or seen someone with BU (P-values <0.001). The same pattern was seen with Christians of the Pentecostal denomination (P-values: <0.001 – 0.023), people having attained higher education (P-values <0.001), salaried workers (P-values <0.001) and singles (P-values<0.001) (Table 2).

**Fig 2.**
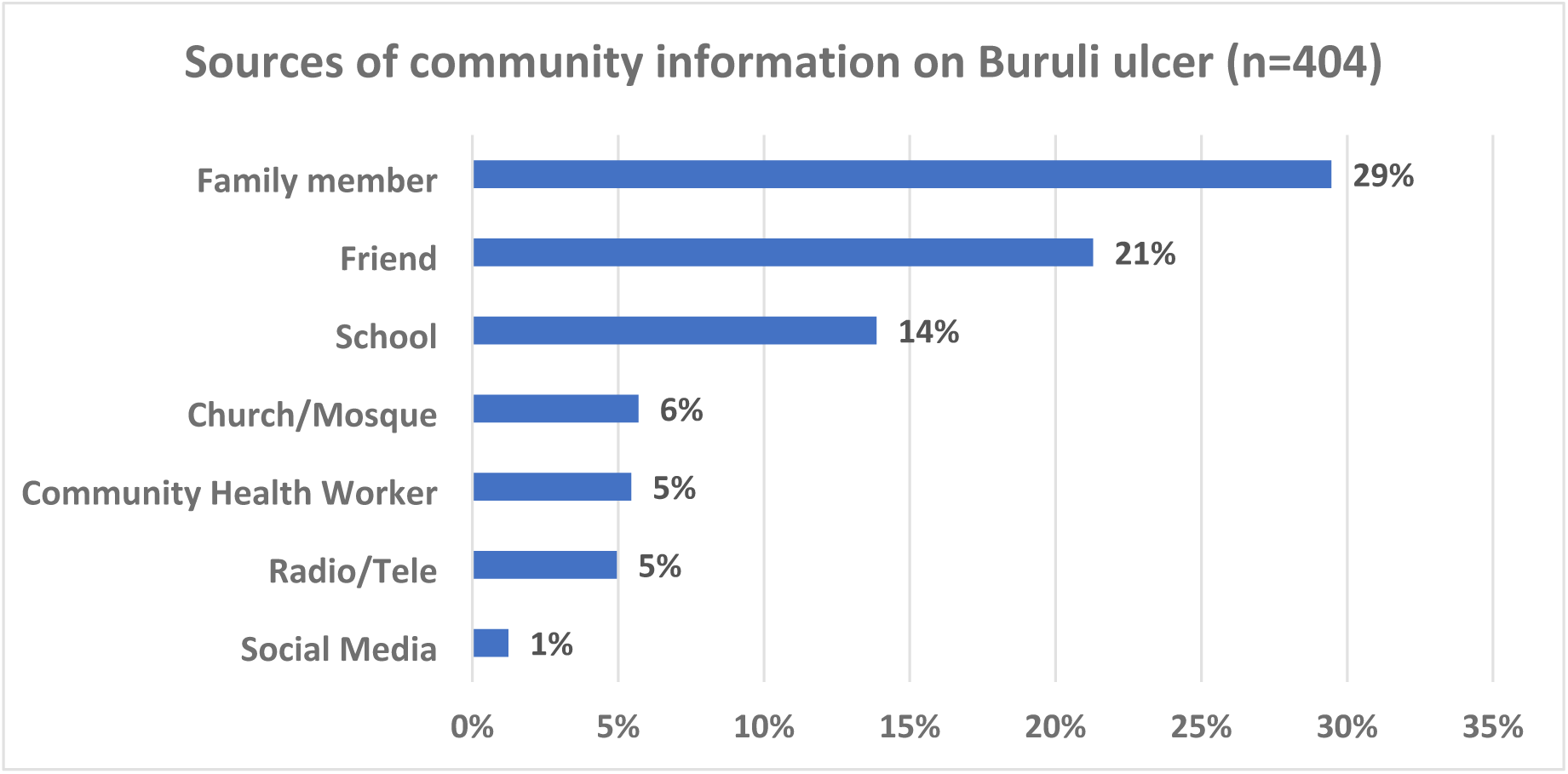
Sources of information on BU.

**Table 2.**
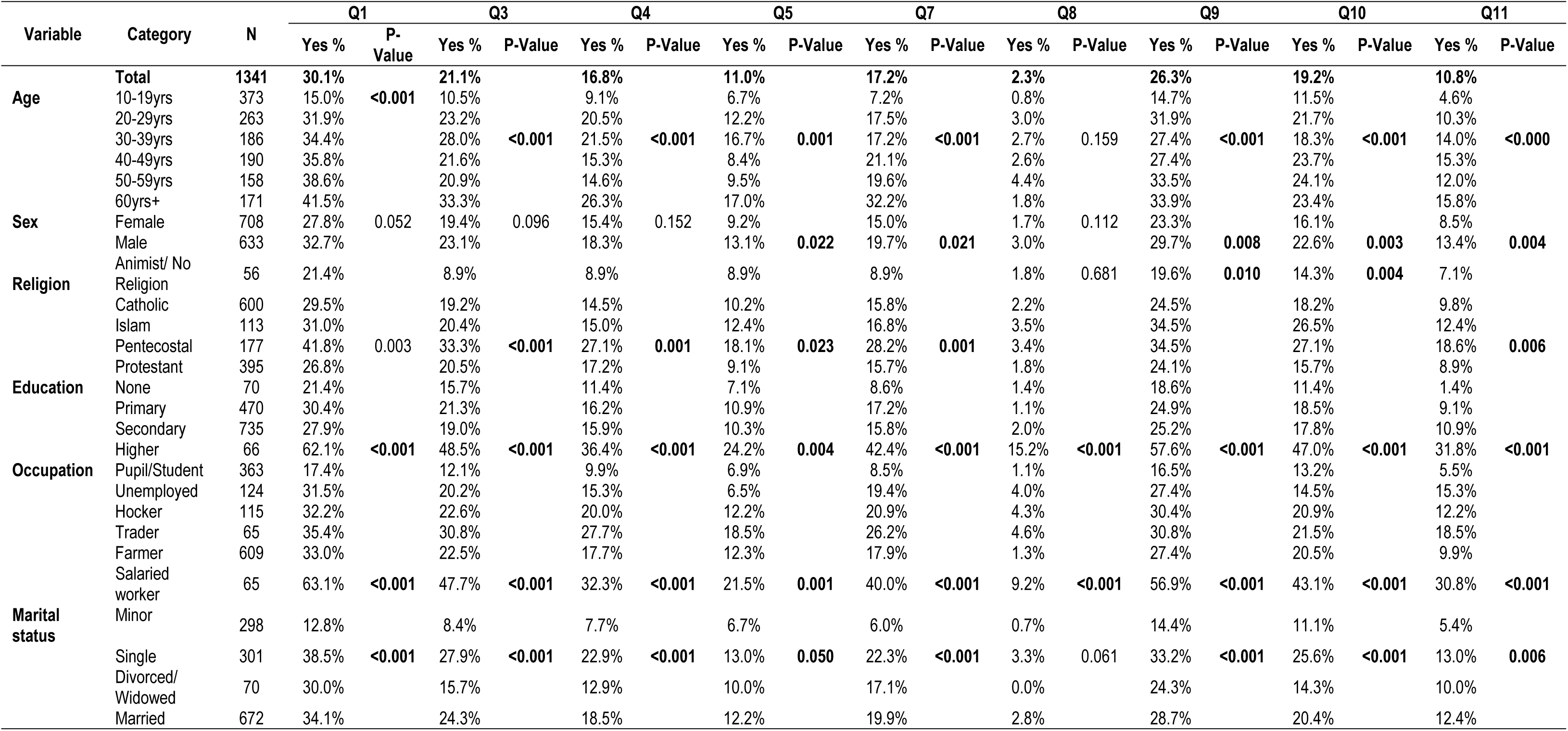
Relationship between socio-demographic variables and knowledge of Buruli ulcer.

Regarding knowledge of clinical manifestations BU, 515(38%) of our respondents attempted to cite, with 231(17.2%) of them citing the correct manifestations of BU including: ulcers(38.1%), oedema(11.3%), nodule(6.8%) and plaque(1.6%) (Table 2 and Fig 3a & b, correct manifestations heighted in red bars). Of the respondents, 451(34%) indicated the cause of BU, and as low as 2.3% knew that BU is caused by a microbe. Majority (54.8%) did not know any cause at all and many cited erroneous causes of BU that were probably based on cultural beliefs and misconceptions (Table 2 and Fig 3c & d). For instance, 9.8% cited poor hygiene, 8.0% cited witchcraft or supernatural forces, 5.3% spontaneous occurrence, 4.9% insect bite, as causes of Buruli ulcer (Fig 3c & d). Regarding knowledge of treatment and prevention of BU, only 19.2% of our respondents believed that BU was curable and a smaller proportion (10.8%) understood its correct preventive measures (Table 2). Of the 449(33.5) of respondents who gave their opinions on where to seek treatment for BU, 78.6% would advise PWBU to visit the hospital for treatment, and 8.2% and 0.9% would prefer traditional healers or prayer homes respective (Fig 4a & b). Elderly persons, those with a secondary or higher level of education and the male gender were more likely to have better Knowledge of manifestations, causes, correct source of treatment and preventive measures of BU among our respondents (P-value <0.05) (Table 2).

**Fig 3.**
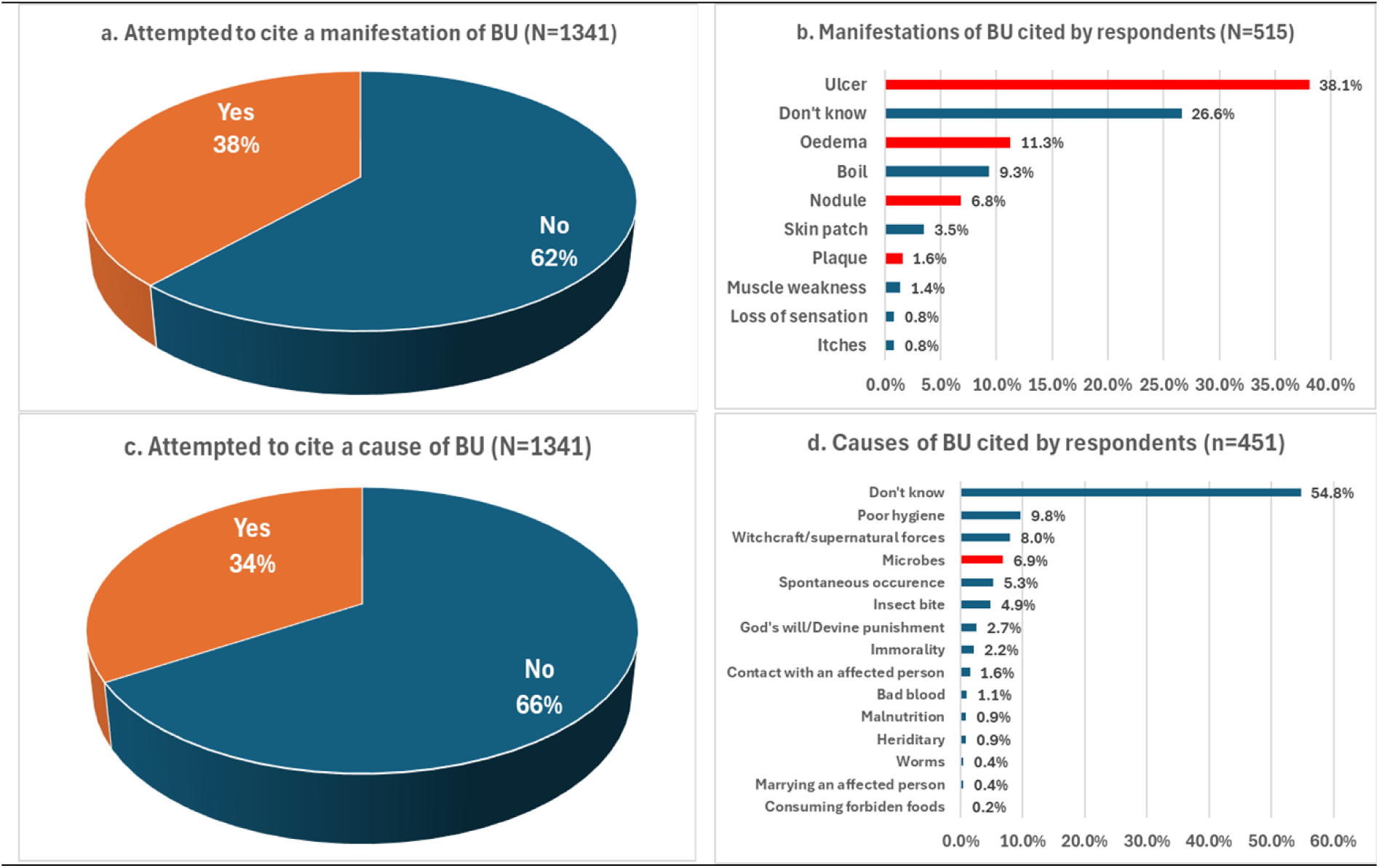
Community knowledge on the causes and manifestations of BU. Thirty-eight percent of respondents attempted to cite the manifestations of BU (Fig 3a), of whom some correctly cited ulcers(38.1%), oedema(11.3%), nodule(6.8%) and plaque(1.6%) highlighted in red (Fig 3b). Equally, 34% attempted to cite the causes of BU (Fig 3c) and only 6.9% of rightly indicated microbes. The rest cited erroneous causes including witchcraft, poor hygiene, insect bite and spontaneous occurrence (Fig 3d).

**Fig 4.**
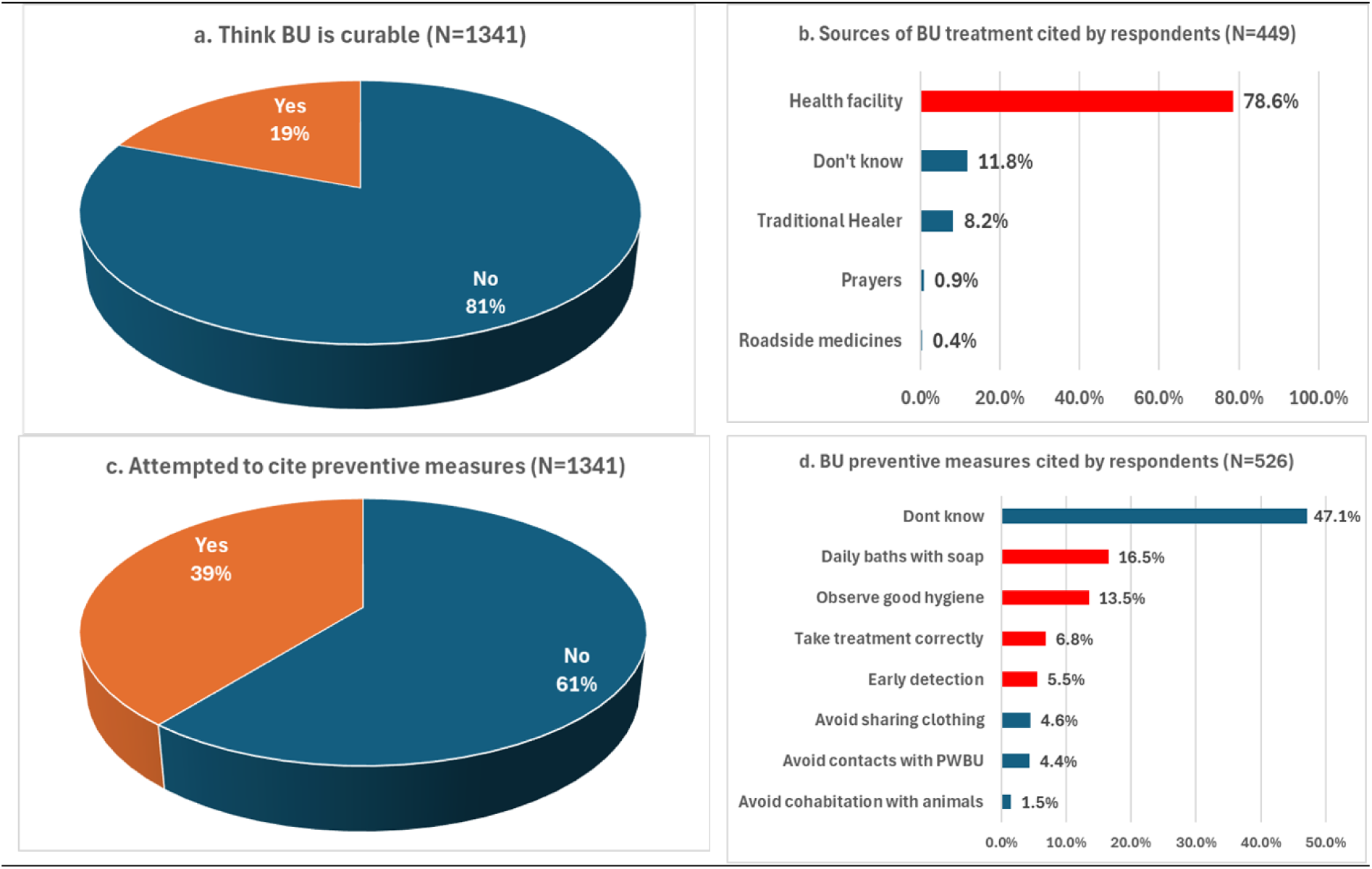
Community knowledge regarding treatment and prevention of BU. Nineteen percent of respondents believed that BU is curable (Fig 4a). Of those who gave opinions on BU treatment source, 78.6% advised health facilities meanwhile, 8.2% and 0.9% preferred traditional healers and prayer homes respectively (Fig 4b). some 39% cited BU preventive measures, of whom 16.5% indicated daily baths with soap, 13.5% good hygiene, 5.5% advising early diagnosis and 6.8% correct treatment, that were deemed plausible and highlighted in red(Fig 4d).

### Attitudes regarding BU

The attitudes of our respondents regarding BU were very poor (Table 3). Only 19.5% of respondents would shake hands and 16.4% would share the same plate with a PWBU; 23.7% would conceal from relatives or friends if they had BU; 14.1% would allow their children play with PWBU and 12.2% would accept their child marry someone who has or had BU. About 17% of our respondents would accept equal employment for PWBU or allow their participation in the community. Only 27.4% held that PWBU should be respected in the community. Attitudes of our respondents regarding BU were influenced by their socio-demographic characteristics as elderly persons, those of the male gender, widows or currently married persons, Christian of the Pentecostal denomination and those who had attained secondary and higher levels of education had better attitudes towards PWBU (P-values <0.05) (Table 3).

**Table 3.**
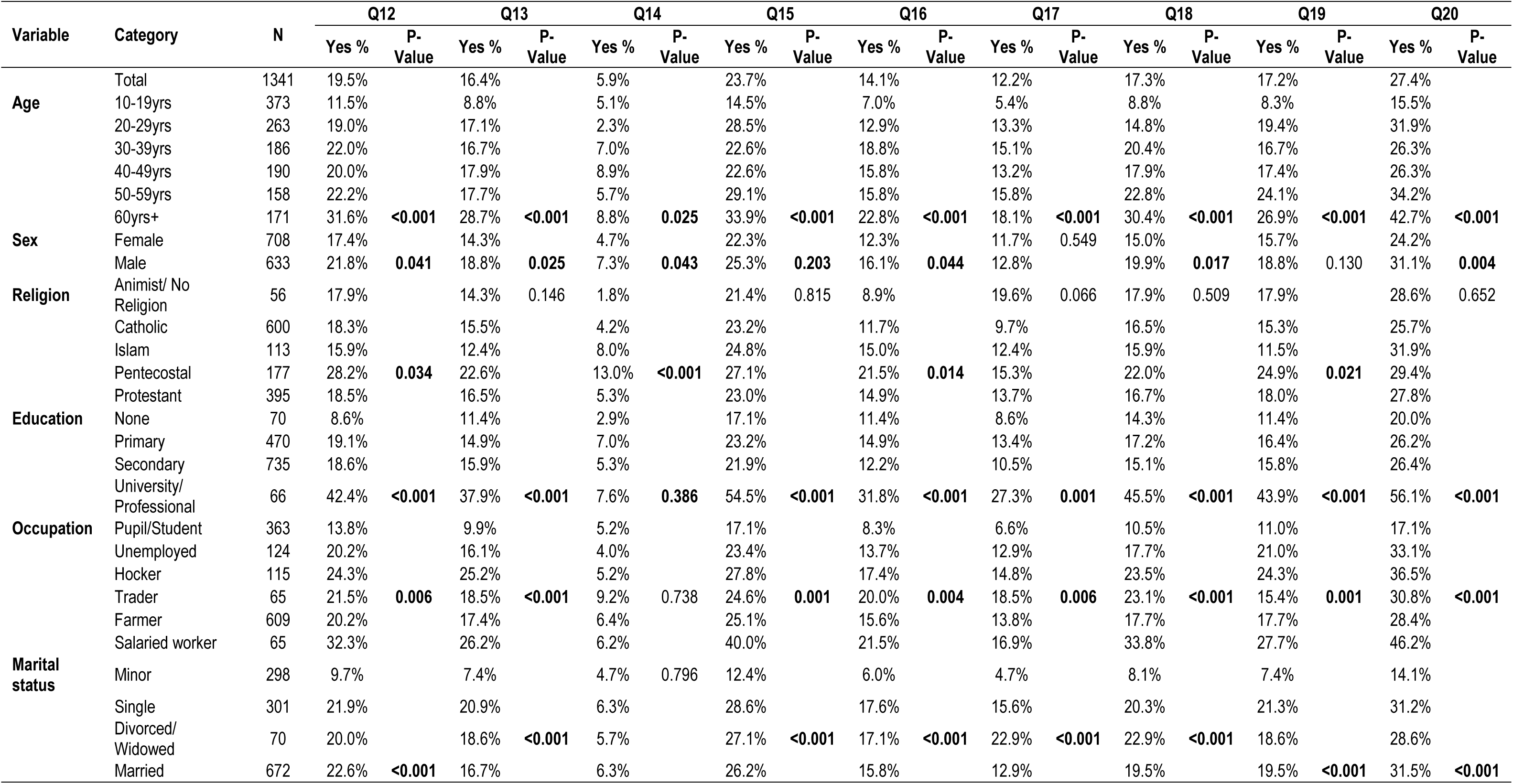
Relationship between socio-demographic variables and attitudes and perceptions regarding Buruli ulcer.

An analysis of the effect of knowledge and perceptions regarding BU on the attitudes of our respondents towards PWBU is shown in table 4. All the attitudes considered were positively influenced by high levels of familiarity with BU, knowledge of the correct cause and clinical manifestations of BU, the understanding that BU is curable, and treatable at the hospital, as well as the knowledge of BU preventive measures (P-values <0.05).

**Table 4.** Relationship between attitudes/perceptions and knowledge of Buruli ulcer.

| Knowledge question | Response | N | Q12 |  | Q13 |  | Q14 |  | Q15 |  | Q16 |  | Q17 |  | Q18 |  | Q19 |  | Q20 |  |
| --- | --- | --- | --- | --- | --- | --- | --- | --- | --- | --- | --- | --- | --- | --- | --- | --- | --- | --- | --- | --- |
|  |  |  | Yes % | P-value | Yes % | P-value | Yes % | P-value | Yes % | P-value | Yes % | P-value | Yes % | P-value | Yes % | P-value | Yes % | P-value | Yes % | P-value |
|  | Total | 1341 | 19.5% |  | 16.4% |  | 5.9% |  | 23.7% |  | 14.1% |  | 12.2% |  | 17.3% |  | 17.2% |  | 27.4% |  |
| Heard about BU | No | 937 | 1.5% |  | 1.2% |  | 0.9% |  | 2.8% |  | 0.6% |  | 0.9% |  | 1.7% |  | 1.7% |  | 5.4% |  |
|  | Yes | 404 | 61.1% | <0.001 | 51.7% | <0.001 | 17.6% | <0.001 | 72.3% | <0.001 | 45.3% | <0.001 | 38.6% | <0.001 | 53.5% | <0.001 | 53.0% | <0.001 | 78.5% | <0.001 |
| Has seen someone with BU | No | 1058 | 6.5% |  | 6.0% |  | 2.5% |  | 9.5% |  | 4.7% |  | 4.5% |  | 6.4% |  | 6.6% |  | 12.9% |  |
|  | Yes | 283 | 67.8% | <0.001 | 55.5% | <0.001 | 18.7% | <0.001 | 76.7% | <0.001 | 49.1% | <0.001 | 41.0% | <0.001 | 58.0% | <0.001 | 56.5% | <0.001 | 81.6% | <0.001 |
| Know someone who had/has BU | No | 1116 | 9.1% | <0.001 | 7.8% | <0.001 | 3.3% | <0.001 | 12.9% | <0.001 | 6.6% | <0.001 | 5.6% | <0.001 | 9.1% | <0.001 | 8.7% | <0.001 | 16.0% | <0.001 |
|  | Yes | 225 | 71.1% |  | 59.1% |  | 18.7% |  | 77.3% |  | 51.1% |  | 44.9% |  | 58.2% |  | 59.1% |  | 84.0% |  |
| Have a relative who suffered BU | No | 1193 | 13.0% |  | 11.1% |  | 3.8% |  | 16.6% |  | 9.3% |  | 8.0% |  | 12.0% |  | 11.6% |  | 20.5% |  |
|  | Yes | 148 | 71.6% | <0.001 | 58.8% | <0.001 | 23.0% | <0.001 | 81.1% | <0.001 | 52.7% | <0.001 | 46.6% | <0.001 | 60.1% | <0.001 | 62.2% | <0.001 | 83.1% | <0.001 |
| Knows how BU manifests | No | 1288 | 16.8% |  | 14.4% |  | 5.3% |  | 21.1% |  | 12.3% |  | 10.9% |  | 15.5% |  | 15.6% |  | 25.0% |  |
|  | Yes | 53 | 84.9% | <0.001 | 64.2% | <0.001 | 20.8% | <0.001 | 86.8% | <0.001 | 58.5% | <0.001 | 43.4% | <0.001 | 60.4% | <0.001 | 54.7% | <0.001 | 86.8% | <0.001 |
| Knows the cause of BU | No | 1310 | 18.2% |  | 15.2% |  | 5.8% |  | 22.5% |  | 13.1% |  | 11.2% |  | 16.1% |  | 16.1% |  | 26.1% |  |
|  | Yes | 31 | 71.0% | <0.001 | 67.7% | <0.001 | 9.7% | 0.365 | 74.2% | <0.001 | 54.8% | <0.001 | 54.8% | <0.001 | 67.7% | <0.001 | 61.3% | <0.001 | 83.9% | <0.001 |
| Know that BU is treatable at the hospital | No | 988 | 5.5% |  | 4.3% |  | 1.2% |  | 6.8% |  | 3.8% |  | 3.1% |  | 4.8% |  | 5.5% |  | 8.6% |  |
|  | Yes | 353 | 58.6% | <0.001 | 50.4% | <0.001 | 19.0% | <0.001 | 71.1% | <0.001 | 42.8% | <0.001 | 37.7% | <0.001 | 52.4% | <0.001 | 49.9% | <0.001 | 80.2% | <0.001 |
| Understands that BU is curable | No | 1084 | 10.1% |  | 8.8% |  | 2.8% |  | 11.9% |  | 6.9% |  | 6.2% |  | 9.1% |  | 9.5% |  | 14.8% |  |
|  | Yes | 257 | 59.1% | <0.001 | 48.6% | <0.001 | 19.1% | <0.001 | 73.5% | <0.001 | 44.4% | <0.001 | 37.7% | <0.001 | 51.8% | <0.001 | 49.4% | <0.001 | 80.9% | <0.001 |
| Knows preventive measures of BU | No | 1284 | 17.8% |  | 14.3% |  | 5.1% |  | 21.7% |  | 12.7% |  | 10.9% |  | 15.5% |  | 15.6% |  | 25.2% |  |
|  | Yes | 57 | 57.9% | <0.001 | 63.2% | <0.001 | 22.8% | <0.001 | 70.2% | <0.001 | 45.6% | <0.001 | 42.1% | <0.001 | 57.9% | <0.001 | 52.6% | <0.001 | 78.9% | <0.001 |

### Problems faced by people with Buruli ulcer (PWBU)

About a quarter (26%) of our respondents admitted that PWBU do face some challenges in society, at a personal level and/or with their families. The challenges included difficulties getting employment (53.2%), interacting with people (58.9%), getting married (27.9%), getting admission into school (15.8%) or even buying food (9.9%). At the level of family, the respondents held that PWBU have problems with their spouses (16.6%), are a source of problems (13.0%) and shame (8.2%) to their families, and the reason why their family members have difficulties getting married (17.7%) (Fig 5).

**Fig 5.**
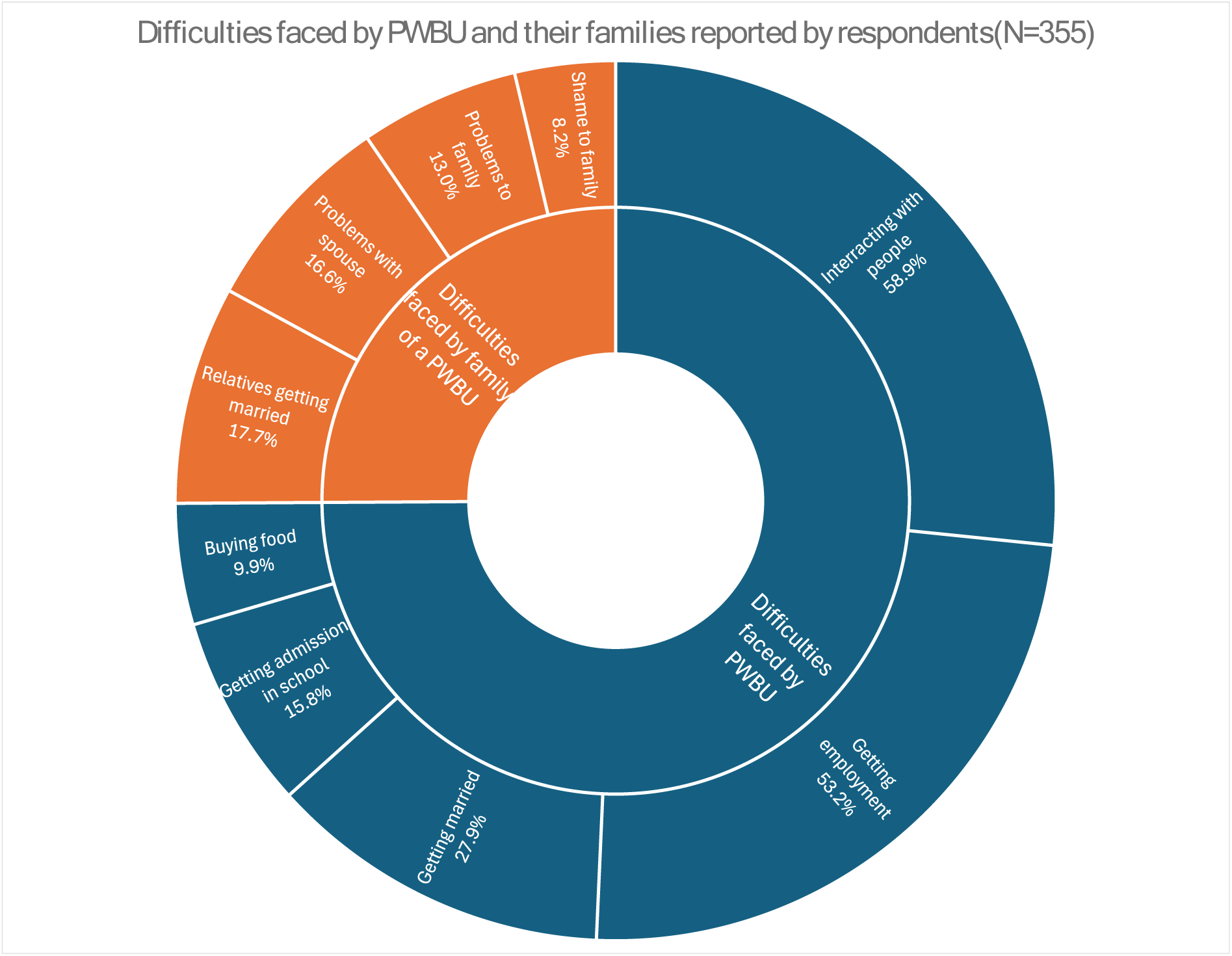
Difficulties faced by people with Buruli ulcer: These difficulties were at two levels. At the individual level where PWBU were perceived to face challenges interacting with people, getting employment, getting married or getting admission to schools. The perceived challenges extended from the individual patient his7her family, bringing shame to family members and problems especially with spouses and with relatives getting married.

### Independent predictors of attitudes towards PWBU

A binary logistic regression analysis inputting socio-demographic variables, familiarity, knowledge and perceptions that influenced attitudes of our respondents as independent variables, and each of the attitudes under evaluation as dependent variable; in a stepwise forward likelihood model was carried out to identify independent predictors attitudes regarding BU. Table 5 shows detail of the outcome. The prominent predictors of positive attitudes identified were: Having heard about BU, knowing that BU is treatable in the hospital, and thinking that BU occurs spontaneously, as they were predictors of 5 to 8 positive attitudes. Equally, the belief that BU is caused by supernatural forces, or poor personal hygiene or staying with a PWBU were identified as predictors of negative attitudes towards PWBU.

**Table 5:** Predictors of attitudes regarding Buruli ulcer.

| Positive attitudes | Predictors | OR | 95%CI |  | P-value |
| --- | --- | --- | --- | --- | --- |
|  |  |  | Lower | Upper |  |
| Would shake hands with someone suffering from Buruli ulcer | Having attained secondary education | 3.92 | 1.17 | 13.16 | 0.027 |
|  | Having attained Higher education | 5.40 | 1.38 | 21.19 | 0.016 |
|  | Having head about BU | 33.14 | 16.61 | 66.12 | <0.001 |
|  | Knowing someone with BU | 2.26 | 1.45 | 3.53 | <0.001 |
|  | Thinks BU occurs spontaneously | 17.82 | 2.32 | 136.96 | 0.006 |
|  | Knowing that BU is treatable in the hospital | 2.72 | 1.68 | 4.41 | <0.001 |
| Would eat from the same plate with someone who has Buruli ulcer | Having head about BU | 40.51 | 19.79 | 82.9 | <0.001 |
|  | Believe BU is caused by supernatural forces | 0.04 | 0.01 | 0.54 | 0.015 |
|  | Thinks BU occurs spontaneously | 2.69 | 1.02 | 7.11 | 0.046 |
|  | Knowing that BU is treatable in the hospital | 2.86 | 1.76 | 4.45 | <0.001 |
| Would not be ashamed of a family member suffering from Buruli ulcer | Being 50yrs old and above | 0.39 | 0.16 | 0.98 | 0.044 |
|  | Being of the Pentecostal Christian denomination | 2.36 | 1.13 | 4.92 | 0.022 |
|  | Having head about BU | 8.65 | 3.31 | 22.6 | <0.001 |
|  | Knowing that BU is treatable in the hospital | 5.54 | 2.46 | 12.51 | <0.001 |
| Would be willing to let a relative/friend know about it if he/she had Buruli ulcer | Being 30-39yrs old | 0.41 | 0.19 | 0.88 | 0.022 |
|  | Being 40-49yrs old | 0.44 | 0.21 | 0.92 | 0.030 |
|  | Having head about BU | 24.72 | 13.98 | 43.69 | <0.001 |
|  | Having a relative with BU | 1.97 | 1.15 | 3.38 | 0.014 |
|  | Knowing that BU is treatable in the hospital | 3.33 | 2.06 | 5.4 | <0.001 |
|  | Thinks BU is curable | 1.78 | 1.11 | 2.85 | 0.017 |
| Would allow his/her child to play with a person affected by Buruli ulcer | Having head about BU | 76.13 | 30.9 | 187.58 | <0.001 |
|  | Thinks BU occurs spontaneously | 2.67 | 1.08 | 6.61 | 0.034 |
|  | Knowing that BU is treatable in the hospital | 1.99 | 1.23 | 3.23 | 0.005 |
| Would accept his/her child marry someone who has/had Buruli ulcer | Being divorced | 5.12 | 1.75 | 14.99 | 0.003 |
|  | Having head about BU | 29.77 | 12.64 | 70.12 | <0.001 |
|  | Knowing someone with BU | 1.76 | 1.14 | 2.71 | 0.011 |
|  | Thinks BU occurs spontaneously | 2.51 | 1.04 | 6.07 | 0.041 |
|  | Thinks BU is caused by living with a PWBU | 0.08 | 0.01 | 0.71 | 0.023 |
|  | Knowing that BU is treatable in the hospital | 2.26 | 1.34 | 3.81 | 0.002 |
| Thinks a person with Buruli ulcer should take part in social activities in the community like anyone else | Having head about BU | 25.99 | 13.97 | 48.36 | <0.001 |
|  | Knowing that BU is treatable in the hospital | 3.07 | 1.93 | 4.87 | <0.001 |
|  | Being 60 yrs or older | 2.34 | 1.18 | 4.67 | 0.015 |
|  | Knowing the correct cause of BU | 3.07 | 1.93 | 4.87 | <0.001 |
| Thinks a person with Buruli ulcer should be employed like anyone else | Having head about BU | 30.74 | 16.89 | 55.95 | <0.001 |
|  | Having a relative with BU | 1.93 | 1.23 | 3.01 | 0.004 |
|  | Thinks BU is caused by supernatural forces | 0.23 | 0.06 | 0.99 | 0.049 |
|  | Thinks BU is due to poor personal hygiene | 0.38 | 0.18 | 0.79 | 0.009 |
| Thinks a person with Buruli ulcer should be respected in the community like anyone else | Having head about BU | 13.08 | 7.94 | 21.56 | <0.001 |
|  | Thinks BU occurs spontaneously | 3.62 | 1.05 | 12.46 | 0.041 |
|  | Knowing that BU is treatable in the hospital | 5.51 | 3.34 | 9.07 | <0.001 |
|  | Thinks BU is curable | 1.82 | 1.08 | 3.08 | 0.026 |

## Discussion

### Knowledge and perceptions regarding BU

This study has revealed low community knowledge and familiarity regarding BU in the Bafia Health District as only 30.1% had heard about the disease, 21.1% knew and 16.8% had seen someone with BU, and 11% declared having a relative with the condition. This contrasts with findings in the Southwest (21) and Centre (37) regions of Cameroon, and Obom district in Ghana(22) where 84.4%, 44.9% and 95.5% of respondents respectively had heard about BU, but comparable with findings in southern Nigeria where 35% of respondents knew of BU (38). The studies in the southwest of Cameroon and Ghana were carried out in known BU endemic health districts where some BU control activities including community awareness had been ongoing, unlike in our study setting in Bafia health district and study areas in southern Nigeria that were newly confirmed for BU and control activities had not begun. The major sources of BU information to our respondents were family members (29%), friends (21%) and schools (16%) rather than health personnel, community health workers (5%) and the media (5%) (Fig 2) often considered as traditional sources of health information. This corroborates with findings by Koka in Ghana where 60% of respondents got their information on BU from community members and only 6.3% and 4.6% got their information from health personnel and the media respectively(22). Having attained secondary education or higher, being elderly or being a male were associated with better knowledge of BU (p-value <0.05) (Table 2). The few respondents (17.2%) with good knowledge of BU manifestations in our study were aware of its early signs and symptoms, citing Ulcer, oedema, nodule and plaque (Fig 3b), which were consistent with clinical forms of the diseases(8,15,16,39). Information on the cause of BU was very unreliable and erroneous for the most part, as majority of respondents did not know the cause at all and some of them attributed BU to witchcraft, immorality, divine punishment, insect bite or spontaneous occurrence (Fig 3d). These findings corroborates with those in the southwest of Cameroon(21) and southern Nigeria(38,40) where majority of respondents did not know the cause of BU and those who attempted cited witchcraft, worms, poor hygiene, insect bites, with very few correctly indicating bacterial infection as cause. The erroneous causes of BU cited by respondents in our study correlates with low familiarity and awareness of BU, but were interestingly consistent with findings in known endemic areas where control activities are being implemented(21,22,28,38). These findings demonstrate deep entrenchment in cultural beliefs and perceptions regarding BU. In Bankim, an endemic health district of Cameroon, BU is said to have four origins, three of them being related to witchcraft, ancestral curse or some inborn supernatural power denoted as *Mgbati* (41,42). In Ayos and Akonolinga, health districts around the Nyong River valley in Cameroon, BU locally known as *atom*, is attributed to a spell or witchcraft affliction following transgression of social norms like stealing from someone’s farm(29). These beliefs have contributed to poor health-seeking behaviors among victims of BU in endemic communities, where a good proportion would prefer traditional healers or prayers as first source of BU treatment(21,28,29,38,42) leading to delays in correct diagnosis and adequate treatment(22,28,29,42).

About 19.2% of our respondents believed that BU was curable, and 78.6% of those who gave their opinion on where to seek BU treatment, would advise victims to go to a health facility, and 8.2% and 0.9% advised traditional healers and prayer homes (Fig 4b) for treatment of BU. These findings are poor compared to those in the southwest of Cameroon, where 87.1% of participants believed that BU was curable, but better than same study when considering treatment-seeking attitudes as 63%, 20% and 17% would respectively refer patients to the hospital, traditional healers and prayers homes(21). The belief in mystical origins of BU held in almost all BU endemic zones across Africa is one of the main drivers for seeking traditional or herbal medicine for BU treatment. This has been demonstrated in Ayos, Akonolinga(29) and Bankim(41) health districts, as well as in the southwest(21) of Cameroon, and also in Ghana(22,28) and southern Nigeria(38). The implications of low knowledge level and poor community perceptions regarding BU as revealed in this study for BU control in Bafia health district is enormous. Even though low, existing community knowledge could be a foundation upon which to develop a successful BU control intervention in the Bafia health district. The national control programme may however need to engage an in-dept anthropological exploration of cultural beliefs and practices regarding BU, and outcomes considered in the development of control strategies adapted to the district as was the case in Bankim health district(41).

### Attitudes towards PWBU

The attitudes of our respondents towards PWBU were generally very negative (Table 3). Less than 20% would shake hands or share from the same plate with a PWBU. This could probably explain why 23.7% of our respondents declared that they would conceal it if they had BU. Similar attitudes were described in southern Nigeria and Ghana where community members would not interact with PWBU for fear of contamination(22,38). Close to 6% of our respondents admitted that they would be ashamed if they had a relative with BU, and 23.7% of them would conceal from relatives and friends if they had BU. These kinds of attitudes obviously take their root in cultural beliefs about the cause of BU (Fig 3d). In the Ayos and Akonolinga health districts of Cameroon where BU *(Atom)* is attributed to transgression of some social norms(29), victims of BU who fear to be tagged would prefer concealment. Our study also revealed that only 14.1% and 12.2% would respectively allow their child to play or marry someone with BU. The perceptions that BU was caused by poor hygiene, heredity, contact with an affected person or marrying an affected person (Fig 3d) could form the basis of these attitudes. Sensitization messages within the framework of a BU control strategy in Bafia health districts must take these into consideration, and package counter information focused on facts about BU including: its natural occurrence, biological etiology, its non-hereditary nature, the non-human-to-human transmission of BU, and its curable nature through early diagnosis and adequate biomedical management(7,43,44).

About 17% of our respondents approved of PWBU participating in normal social activities or being given equal employment like anyone else. In these aspects, attitudes towards PWBU in this study were worse compared to those reported in the south west of Cameroon and in southern Nigeria, where higher proportions of respondents would allow free interaction of PWBU as well as equal employment for them(21,38). Only 27.4% of our respondents felt that PWBU should be respected in the community. These depict the high level of community stigma PWBU could face in Bafia health district. In endemic zones where BU control activities are being implemented, community attitudes towards PWBU tend to be much more improved(21,38,45). In this study, community attitudes regarding BU were influenced by sociodemographic variables (Table 3) as well as knowledge of and familiarity with BU (P<0.05) (Table 4). The male gender, being elderly, having attained at least primary level of education, and being of the Pentecostal Christian denomination were more likely to portray positive attitudes towards PWBU (P<0.05).

Answering the question on what problems PWBU face, our respondents cited a litany of problems that could be grouped into individual level and family level problems (Fig 5). More than half of those who responded to this question indicated that PWBU face difficulties interacting with people in society or getting employment, and over a quarter said they face challenges getting married. These difficulties extended beyond the individual patients to affecting their families as respondents indicated that family members of PWBU do face problems including feeling ashamed, difficulties getting married themselves (Fig 5). Of course, these difficulties faced by PWBU correlate with community attitudes towards them. The consequences on PWBU would be self-stigma manifested by concealment, self-isolation, and delay in seeking treatment (22,46).

Our study evaluated nine attitudes altogether. Three to six independent predictors were identified for each of the nine attitudes. The beliefs that BU is caused by supernatural forces, or by living with someone with BU, or by poor personal hygiene, or being 30 years of age and older, were predictors of negative attitudes (Table 5). On the other hand: having heard of BU, knowing someone with BU, having a relative with BU, the beliefs that BU is curable and treatable at the hospital, and that BU occurs spontaneously, having attained at least secondary education, being a Christian, were identified as predictors of positive attitudes (Table 5). The implication of public health here is that a BU control intervention in Bafia health district with a strong component of education for behavior change that focuses on enhancing community knowledge on BU and using schools and churches and channels of communication could ensure its effectiveness.

### Conclusion and recommendations

We conclude that familiarity with and knowledge of BU were low in Bafia health district. The major sources of BU information were family, friends and schools. The community knowledge of BU was marked by erroneous perceptions regarding its cause, manifestations. A relatively low proportion of respondents understood that BU is curable, of whom more than 2/3^rd^ advised treatment at health facility. Attitudes regarding BU were equally poor among our respondents. Negative attitudes were independently associated with the beliefs that BU is caused by supernatural forces, or by living with someone with BU, or by poor personal hygiene, or being 30 years of age and older, meanwhile positive attitudes were driven by: having heard of BU, knowing someone with BU, having a relative with BU, the beliefs that BU is curable and treatable at the hospital, and that BU occurs spontaneously, having attained at least secondary education, being a Christian.

We recommend that a BU control intervention in Bafia health district must embed a strong component of education for behavior change that focuses on enhancing community knowledge on BU and using schools and churches and channels of communication, with information on: its natural occurrence, biological etiology, its non-hereditary nature, the non-human-to-human transmission of BU, and its curable nature through early diagnosis and adequate biomedical management. This could improve community knowledge and attitudes towards PWBU and ultimately lead to an effective and efficacious BU control intervention in the district.

## Data Availability

All relevant data are within the manuscript and its Supporting Information files

## Declarations

## Disclaimer (Artificial Intelligence)

Author(s) hereby declares that NO generative AI technologies such as Large Language Models (ChatGPT, COPILOT, etc) and text-to-image generators have been used during writing or editing of this manuscript.

## Author contributions

➢ **Conception:** Earnest Njih Tabah,
➢ **Data curation:** Earnest Njih Tabah, Djam Chefor Alain, Njih Irine Ngani Nformi, Loic Douanla Pagning, Nzoyem Tsago Colin, Baran A Bidias Elisabeth, Kouayep Watat Christian Elvis
➢ **Methodology:** Earnest Njih Tabah, Djam Chefor Alain, Njih Irine Ngani Nformi, Loic Douanla Pagning, Nzoyem Tsago Colin, Baran A Bidias Elisabeth, Wanda Franck Eric, Larraitz Ventoso
➢ **Data analysis:** Earnest Njih Tabah, Djam Chefor Alain, Njih Irine Ngani Nformi, Loic Douanla Pagning, Nzoyem Tsago Colin
➢ **Writing the original draft:** Earnest Njih Tabah, Djam Chefor Alain, Njih Irine Ngani Nformi, Loic Douanla Pagning
➢ **Review and editing of manuscripts:** Earnest Njih Tabah, Djam Chefor Alain, Njih Irine Ngani Nformi, Loic Douanla Pagning, Nzoyem Tsago Colin, Baran A Bidias Elisabeth, Kouayep Watat Christian Elvis, Wanda Franck Eric, Larraitz Ventoso

## Acknowledgement

We acknowledge the Bafia health district management team for administrative facilitation for the conduct of the survey.

